# Current Stroke Clinical Outcomes of Direct Anticoagulation Therapy in Patients with Valvular Atrial Fibrillation: A Scoping Review

**DOI:** 10.1101/2023.02.25.23286281

**Authors:** Nisha Dabhi, Lucy Chu, Necrisha Roach, Sherita Chapman

## Abstract

While direct oral anticoagulants (DOACs) have been proven effective in stroke prevention in patients with non-valvular AF (NVAF), there is limited evidence-based guidance on DOAC use in valvular atrial fibrillation. This scoping review aims to examine clinical cerebrovascular outcomes and safety profiles within valvular AF (VAF) patients treated with DOACs. This scoping review was conducted following the Joanna Briggs Institute methodology for scoping reviews and reported consulting the Preferred Reporting Items for Systematic Review and Meta-Analysis extension for scoping reviews checklist. Pubmed, OVID Medline and Web of Science databases were searched for manuscripts published from inception to July 25, 2022 and included based on pre-registered criteria. Data extraction included patient demographics, valvular disease and Afib classification, DOAC type, cerebro-cardiovascular outcomes variables, and bleeding risks. There were 12 studies with 69,741 VAF patients treated with DOACs. At the 12-month follow-up, acute ischemic stroke and major bleeding events occurred in 3.5% and 5.8% of these patients, respectively. DOACs in VAF were associated with similar clinical cerebrovascular outcomes with decreased risk of fatal bleeding rates when compared to warfarin. More controlled studies are needed to further assess the safety and efficacy of DOACs in this population.

## Introduction

Atrial fibrillation (AF) is the most common type of cardiac arrhythmia, occurring in nearly 8-10% of persons above the age of 80 years.^1,2^ Importantly, AF is associated with a 5-fold increased risk for stroke, and it’s responsible for causing at least 15% of all acute ischemic strokes (AIS) with a higher associated risk of fatal outcome.^1,2^ AF-related strokes have been found to result in larger areas of brain infarction, which subsequently increased risk of disability and mortality compared with strokes of other mechanisms.^3^

Oral anticoagulation (OAC) therapy has been shown to reduce the risk of stroke by 64% in patients with AF.^4^ As such, direct oral anticoagulants (DOACs) are currently recommended as first-line therapy for patients with non-valvular atrial fibrillation, having demonstrated decreased need for regular monitoring (ie. less drug-drug and drug-food interactions), as well as significant reductions in intracranial hemorrhage and fatal bleeding when compared with warfarin.^5^ However, there is limited evidence to guide recommendations on the use of DOACs in valvular atrial fibrillation (ie. atrial fibrillation in patients with prosthetic valves or aortic/mitral valve dysfunction), although the risk of AIS in these patients are up to 17-fold higher than the general population.^6^ This scoping review aims (1) to examine and map the clinical cerebrovascular outcomes and (2) to identify knowledge gaps in patients with valvular AF treated with DOACs.

## Methods

### Literature Search and Review

A scoping review of the literature was performed following the Joanna Briggs Institute (JBI) Methodology for JBI Scoping Reviews and consulted the Preferred Reporting Items for Systematic Reviews and Meta-Analyses (PRISMA) Extension for Scoping Reviews checklist for reporting (Supplement Table 1). The protocol is registered with Open Science Framework. Full inclusion and exclusion screening criteria are presented in **Table 1**. Valvular atrial fibrillation was defined as atrial fibrillation in the presence of aortic stenosis, aortic regurgitation, mitral stenosis, mitral regurgitation with or without bioprosthetic valve repair (ie. patients with mechanical valves were not included).

**Table 1.** Inclusion and Exclusion Criteria for Study

| Inclusion | Exclusion |
| --- | --- |
| (1) Use of DOAC for primary or secondary stroke prevention<br>(2) Diagnosis of atrial fibrillation<br>(3) Valvular atrial fibrillation<br>(4) Provision of outcomes data (i.e. vascular complications, stroke incidence) for patients meeting above criteria | (1) DOACs not used for primary or secondary stroke prevention<br>(2) No diagnosis of atrial fibrillation<br>(3) Non-valvular atrial fibrillation<br>(4) Mechanical heart valves<br>(4) No outcomes data (ie. vascular complications, stroke incidence) |

DOAC: direct oral anticoagulant

The electronic databases Pubmed, Web of Science, and Ovid Medline were searched from inception to July 25, 2022. Keyword search using Boolean operators OR & AND with terms including but not limited to “direct oral anticoagulation,” “valvular,” and “atrial fibrillation” was conducted (Supplement Search Terms). Duplicates of search results from different databases were identified and removed. Two independent, blinded authors (ND, LC) initially screened manuscript titles and abstracts. Following title and abstract review, two authors (ND, LC) evaluated the subsequent full-text manuscripts for inclusion in the final review. Any disagreements following full-text evaluation were discussed between authors. A third author (SC) was available for consultation if no consensus could be reached. Reference lists of selected articles and excluded, relevant reviews of literature were examined to detect articles potentially missed by initial search.

The objectives, inclusion and exclusion criteria, and methods for this scoping review were specified in advance and documented in a protocol.

### Data Extraction

Data from all included full-text manuscripts were collected within a data extraction spreadsheet by the first author (ND) and then verified by the second author (LC). Any inconsistencies were resolved by consensus. The following baseline data was collected from each included study: age, sex, CHA_2_DS_2_-VASc score, HAS-BLED score, history of valve repair/replacement, histories of prior ischemic stroke, diabetes, and hypertension. Type of valvular disease (ie. aortic, mitral, both) and type of atrial fibrillation (ie. incidental unspecified, paroxysmal, persistent, permanent) of patients in each study were also collected. The following clinical outcomes related data were of interest: type of DOAC, clinical follow-up time, incidences of stroke, myocardial infarction (MI), systemic embolism, gastrointestinal (GI) bleeding, non-major bleeding, major bleeding, and overall mortality. Major bleeding was defined as (1) fatal bleeding and/or, (2) symptomatic bleeding in a critical area or organ (e.g. intracranial) or, (3) bleeding causing a fall in hemoglobin level of 20 g/L or more. The type of study, inclusion criteria, and exclusion criteria of each study were also recorded.

The primary outcomes were rates of AIS and major bleeding in patients with valvular AF on DOAC. Secondary outcomes included rate of non-major bleeding and mortality in patients with valvular AF on DOAC.

### Data Synthesis and Quality Assessment

For each included article, continuous variables such as age, CHA_2_DS_2_-VASc score, and HAS-BLED score were reported either as mean values with standard deviations or median values with interquartile ranges. A crude estimate was computed as the mean of all included studies. Categorical variables (ie. past medical history, type of valvular disease, complications) were reported as proportions for each article. The crude estimate was the total proportion of all included studies. Quality assessment was assessed through the Newcastle-Ottawa Scale (Supplement Table 1).

## Results

There were 12 studies from 2015 to 2022 that included baseline characteristics, complications, or outcomes related data pertaining to patients with VAF.^7-18^ The PRISMA flow diagram detailing the results of the literature search and review is presented in **Figure 1**. Among these 12 studies, five were randomized control trials (RCT), five were retrospective cohort studies, one was a prospective cohort study, and one was a descriptive cohort study (**Table 2**).

**Table 2.**
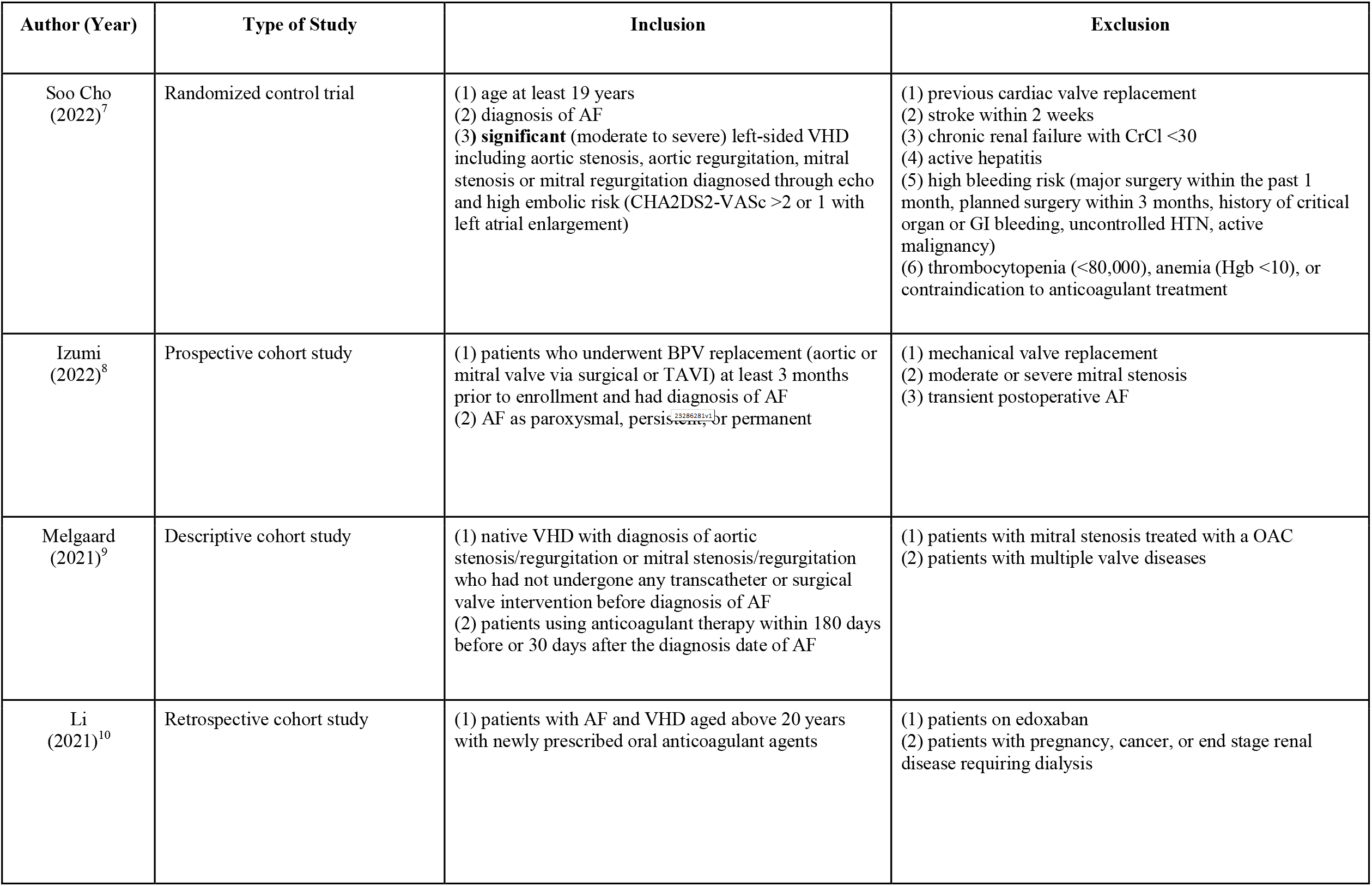
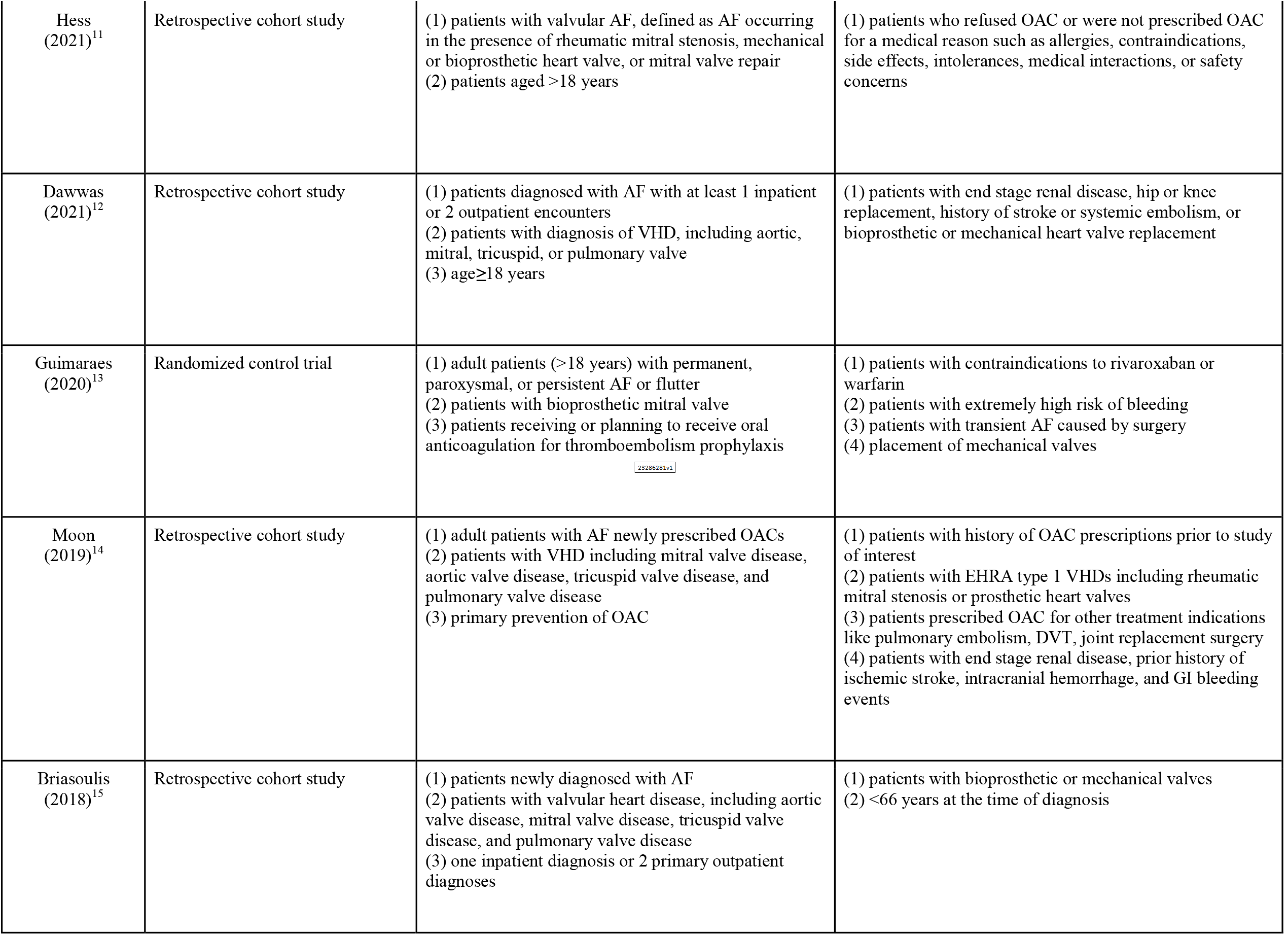
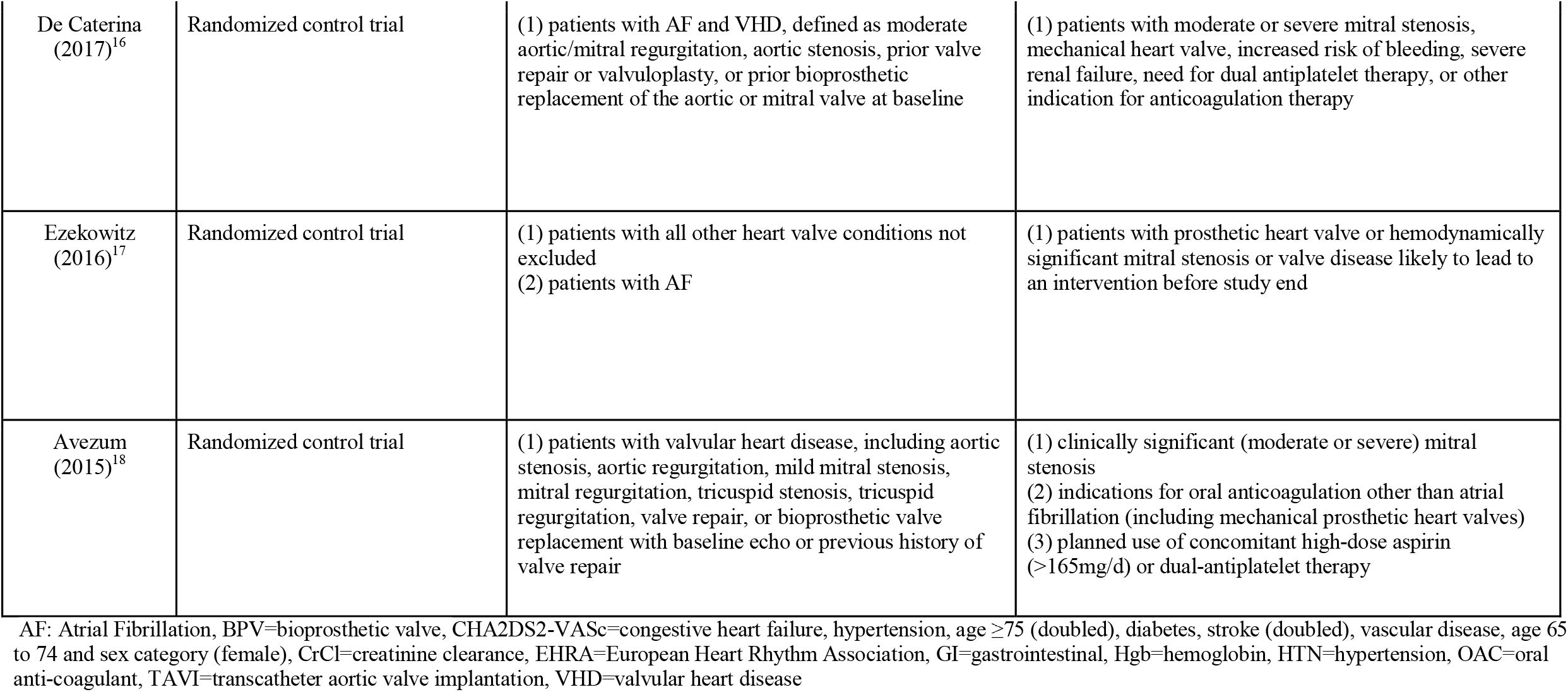
Characteristics of Included Studies

| Author (Year) | Type of Study | Inclusion | Exclusion |
| --- | --- | --- | --- |
| Soo Cho<br>(2022)<sup>7</sup> | Randomized control trial | (1) age at least 19 years<br>(2) diagnosis of AF<br>(3) **significant** (moderate to severe) left-sided VHD including aortic stenosis, aortic regurgitation, mitral stenosis or mitral regurgitation diagnosed through echo and high embolic risk (CHA2DS2-VASc >2 or 1 with left atrial enlargement) | (1) previous cardiac valve replacement<br>(2) stroke within 2 weeks<br>(3) chronic renal failure with CrCl <30<br>(4) active hepatitis<br>(5) high bleeding risk (major surgery within the past 1 month, planned surgery within 3 months, history of critical organ or GI bleeding, uncontrolled HTN, active malignancy)<br>(6) thrombocytopenia (<80,000), anemia (Hgb <10), or contraindication to anticoagulant treatment |
| Izumi<br>(2022)<sup>8</sup> | Prospective cohort study | (1) patients who underwent BPV replacement (aortic or mitral valve via surgical or TAVI) at least 3 months prior to enrollment and had diagnosis of AF<br>(2) AF as paroxysmal, persistent, or permanent | (1) mechanical valve replacement<br>(2) moderate or severe mitral stenosis<br>(3) transient postoperative AF |
| Melgaard<br>(2021)<sup>9</sup> | Descriptive cohort study | (1) native VHD with diagnosis of aortic stenosis/regurgitation or mitral stenosis/regurgitation who had not undergone any transcatheter or surgical valve intervention before diagnosis of AF<br>(2) patients using anticoagulant therapy within 180 days before or 30 days after the diagnosis date of AF | (1) patients with mitral stenosis treated with a OAC<br>(2) patients with multiple valve diseases |
| Li<br>(2021)<sup>10</sup> | Retrospective cohort study | (1) patients with AF and VHD aged above 20 years with newly prescribed oral anticoagulant agents | (1) patients on edoxaban<br>(2) patients with pregnancy, cancer, or end stage renal disease requiring dialysis |
| Hess<br>(2021)<sup>11</sup> | Retrospective cohort study | (1) patients with valvular AF, defined as AF occurring in the presence of rheumatic mitral stenosis, mechanical or bioprosthetic heart valve, or mitral valve repair<br>(2) patients aged >18 years | (1) patients who refused OAC or were not prescribed OAC for a medical reason such as allergies, contraindications, side effects, intolerances, medical interactions, or safety concerns |
| Dawwas<br>(2021)<sup>12</sup> | Retrospective cohort study | (1) patients diagnosed with AF with at least 1 inpatient or 2 outpatient encounters<br>(2) patients with diagnosis of VHD, including aortic, mitral, tricuspid, or pulmonary valve<br>(3) age ≥ 18 years | (1) patients with end stage renal disease, hip or knee replacement, history of stroke or systemic embolism, or bioprosthetic or mechanical heart valve replacement |
| Guimaraes<br>(2020)<sup>13</sup> | Randomized control trial | (1) adult patients (>18 years) with permanent, paroxysmal, or persistent AF or flutter<br>(2) patients with bioprosthetic mitral valve<br>(3) patients receiving or planning to receive oral anticoagulation for thromboembolism prophylaxis | (1) patients with contraindications to rivaroxaban or warfarin<br>(2) patients with extremely high risk of bleeding<br>(3) patients with transient AF caused by surgery<br>(4) placement of mechanical valves |
| Moon<br>(2019)<sup>14</sup> | Retrospective cohort study | (1) adult patients with AF newly prescribed OACs<br>(2) patients with VHD including mitral valve disease, aortic valve disease, tricuspid valve disease, and pulmonary valve disease<br>(3) primary prevention of OAC | (1) patients with history of OAC prescriptions prior to study of interest<br>(2) patients with EHRA type 1 VHDs including rheumatic mitral stenosis or prosthetic heart valves<br>(3) patients prescribed OAC for other treatment indications like pulmonary embolism, DVT, joint replacement surgery<br>(4) patients with end stage renal disease, prior history of ischemic stroke, intracranial hemorrhage, and GI bleeding events |
| Briasoulis<br>(2018)<sup>15</sup> | Retrospective cohort study | (1) patients newly diagnosed with AF<br>(2) patients with valvular heart disease, including aortic valve disease, mitral valve disease, tricuspid valve disease, and pulmonary valve disease<br>(3) one inpatient diagnosis or 2 primary outpatient diagnoses | (1) patients with bioprosthetic or mechanical valves<br>(2) <66 years at the time of diagnosis |
| De Caterina<br>(2017)<sup>16</sup> | Randomized control trial | (1) patients with AF and VHD, defined as moderate aortic/mitral regurgitation, aortic stenosis, prior valve repair or valvuloplasty, or prior bioprosthetic replacement of the aortic or mitral valve at baseline | (1) patients with moderate or severe mitral stenosis, mechanical heart valve, increased risk of bleeding, severe renal failure, need for dual antiplatelet therapy, or other indication for anticoagulation therapy |
| Ezekowitz<br>(2016)<sup>17</sup> | Randomized control trial | (1) patients with all other heart valve conditions not excluded<br>(2) patients with AF | (1) patients with prosthetic heart valve or hemodynamically significant mitral stenosis or valve disease likely to lead to an intervention before study end |
| Avezum<br>(2015)<sup>18</sup> | Randomized control trial | (1) patients with valvular heart disease, including aortic stenosis, aortic regurgitation, mild mitral stenosis, mitral regurgitation, tricuspid stenosis, tricuspid regurgitation, valve repair, or bioprosthetic valve replacement with baseline echo or previous history of valve repair | (1) clinically significant (moderate or severe) mitral stenosis<br>(2) indications for oral anticoagulation other than atrial fibrillation (including mechanical prosthetic heart valves)<br>(3) planned use of concomitant high-dose aspirin (>165mg/d) or dual-antiplatelet therapy |

AF: Atrial Fibrillation, BPV=bioprosthetic valve, CHA2DS2-VASc=congestive heart failure, hypertension, age $\geq 75$ (doubled), diabetes, stroke (doubled), vascular disease, age 65 to 74 and sex category (female), CrCl=creatinine clearance, EHRA=European Heart Rhythm Association, GI=gastrointestinal, Hgb=hemoglobin, HTN=hypertension, OAC=oral anti-coagulant, TAVI=transcatheter aortic valve implantation, VHD=valvular heart disease

**Figure 1.**
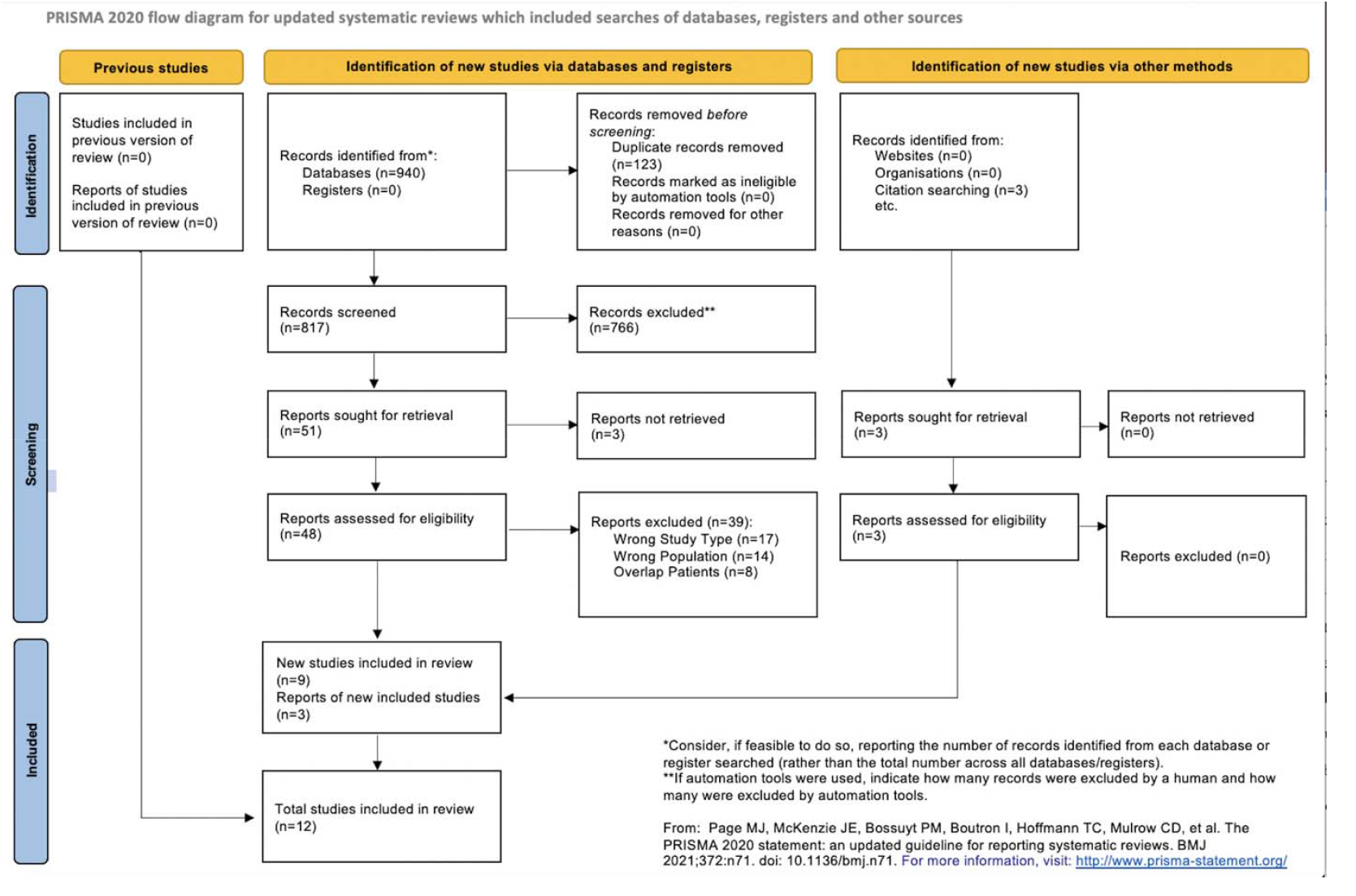
Flow diagram detailing the study selection process

### Demographic Characteristics

**Table 3 and 4** detail the demographic characteristics of the included studies. There were a total of 69,741 VAF patients [female (48%), mean age (73)] treated with DOAC in the 12 included studies. The CHA_2_DS_2_-VASc score was specified in nine studies (36,015 patients), the mean of which was 3.7. Five studies (13,426 patients) noted a HAS-BLED score, of which the mean was 2.3.

**Table 3.**
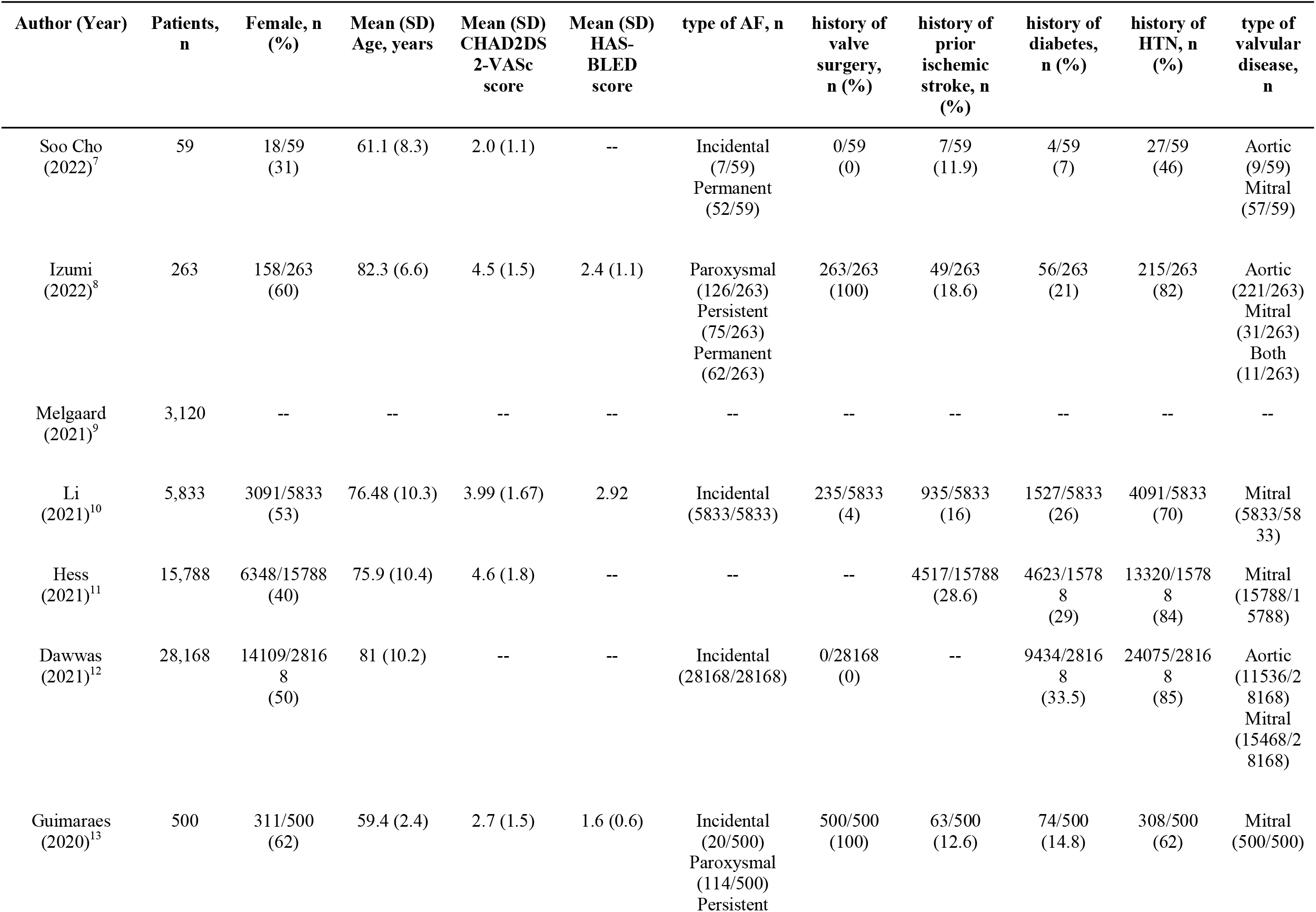
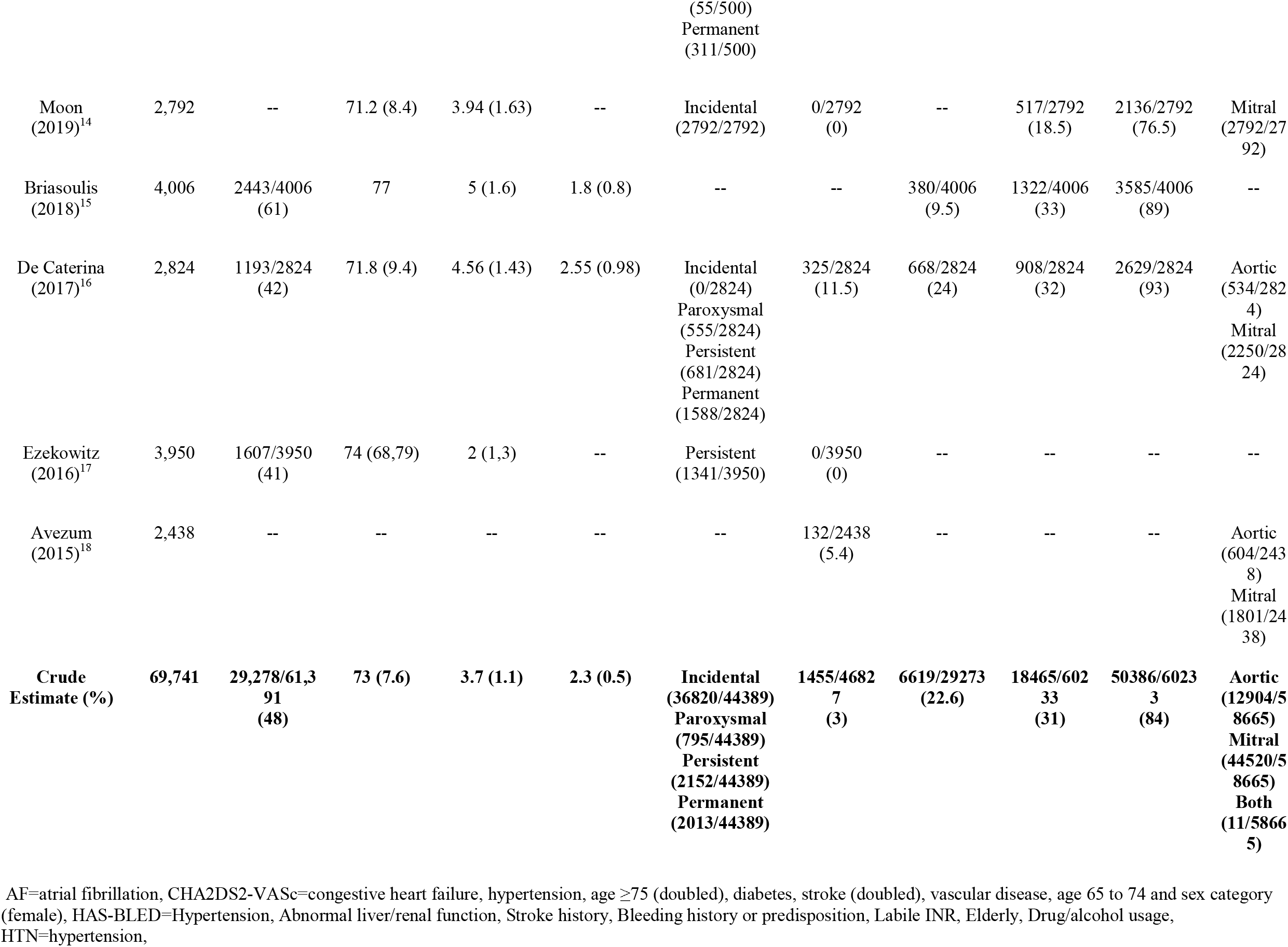
Demographic Characteristics of Patients with Valvular Atrial Fibrillation on DOACs

**Table 4.** Clinical Outcomes in Valvular Atrial Fibrillation Patients on DOACs

| Author (Year) | Type of DOAC | Clinical follow-up, months | Acute ischemic stroke, n (%) | Systemic embolism, n (%) | Major Bleeding, n (%) | Intracranial bleeding, n (%) | GI bleeding, n (%) | Fatal Bleeding, n (%) | Non-major Bleeding, n (%) | Mortality, n (%) |
| --- | --- | --- | --- | --- | --- | --- | --- | --- | --- | --- |
| Soo Cho (2022)<sup>7</sup> | dabigatran | 12 | 0/59<br>(0) | 0/59<br>(0) | 0/59<br>(0) | 0/59<br>(0) | 0/59<br>(0) | 0/59<br>(0) | 3/59<br>(5.1) | 0/59<br>(0) |
| Izumi (2022)<sup>8</sup> | not reported | 12 | 4/263<br>(1.5) | 1/263<br>(0.4) | 7/263<br>(2.7) | 3/263<br>(1.1) | -- | 1/263<br>(0.4) | 14/263<br>(5.3) | 11/263<br>(4.2) |
| Melgaard (2021)<sup>9</sup> | not reported | 12 | -- | 69/3120<br>(2.2) | 97/3120<br>(3.1) | 19/3120<br>(0.6) | 25/3120<br>(0.8) | -- | -- | 617/3120<br>(19.8) |
| Li (2021)<sup>10</sup> | dabigatran,<br>rivaroxaban | 12 | 414/5833<br>(7) | 14/5833<br>(0.2) | 359/5833<br>(6.2) | 74/5833<br>(1.3) | 285/5833<br>(4.9) | -- | -- | 365/5833<br>(6.3) |
| Hess (2021)<sup>11</sup> | dabigatran,<br>rivaroxaban,<br>apixaban,<br>edoxaban | -- | -- | -- | -- | -- | -- | -- | -- | -- |
| Dawwas (2021)<sup>12</sup> | dabigatran,<br>rivaroxaban,<br>apixaban,<br>edoxaban | 12 | 787/28168<br>(3) | -- | 1465/28168<br>(5.2) | 298/28168<br>(1.1) | 1167/28168<br>(4.1) | -- | -- | -- |
| Guimaraes (2020)<sup>13</sup> | rivaroxaban | 12 | 3/500<br>(0.6) | 0/500<br>(0) | 7/500<br>(1.4) | 0/500<br>(0) | -- | 0/500<br>(0) | 61/500<br>(12) | 20/500<br>(4) |
| Moon (2019)<sup>14</sup> | dabigatran,<br>rivaroxaban,<br>apixaban,<br>edoxaban | 12 | 69/2792<br>(2.5) | -- | 75/2792<br>(2.7) | 32/2792<br>(1.1) | 43/2792<br>(1.5) | -- | -- | 161/2792<br>(5.8) |
| Briasoulis (2018)<sup>15</sup> | rivaroxaban,<br>dabigatran | 12 | -- | -- | 167/4006<br>(4.2) | -- | 154/4006<br>(3.8) | -- | -- | 64/4006<br>(1.6) |
| De Caterina<br>(2017)<sup>16</sup> | edoxaban | 33.6 | 103/2824<br>(3.6) | 9/2824<br>(0.3) | 188/2824<br>(6.7) | 28/2824<br>(1.0) | 93/2824<br>(3.3) | 15/2824<br>(0.5) | 257/2824<br>(9.1) | 455/2824<br>(16) |
| Ezekowitz<br>(2016)<sup>17</sup> | dabigatran | 12 | -- | -- | 341/3950<br>(8.6) | 40/3950<br>(1) | -- | -- | -- | 348/3950<br>(8.8) |
| Avezum<br>(2015)<sup>18</sup> | apixaban | 12 | 130/2438<br>(1.2) | -- | 404/2438<br>(4.6) | 31/2438<br>(0.34) | -- | -- | -- | 379/2438<br>(3.7) |
| **Crude Estimate (%)** | **--** | **13.9 (6.5)** | **1510/42877**<br>**(3.5)** | **93/12599**<br>**(0.7)** | **3110/53953**<br>**(5.8)** | **534/49947**<br>**(1.1)** | **1767/46802**<br>**(3.8)** | **16/3646**<br>**(0.4)** | **335/3646**<br>**(9.2)** | **2420/25785**<br>**(9.4)** |

DOAC=direct oral anti-coagulant, GI=gastrointestinal

Regarding comorbid medical conditions, nine studies with 60,233 total patients reported past histories of hypertension or diabetes, which were present in 80% (48327/60233) and 30% (17967/60233) patients, respectively. There were 22.6% (6619/29273) with a past history of ischemic stroke (7 studies).

Ten studies noted the type of DOAC used for patients with VAF (**Table 4**). A combination of either dabigatran, rivaroxaban, apixaban, and edoxaban were prescribed. In five studies, one DOAC was prescribed [dabigatran (2), rivaroxaban (1), edoxaban (1), apixaban (1)].^7,13,16,17,18^ Five studies included patients on multiple DOACs.

### Valvular Disease Characteristics

The type of valvular disease (ie. mitral, aortic, both) was specified in nine studies for 58,665 patients (**Table 3**). Overall, aortic valve stenosis or regurgitation was the primary valvular pathology in 22% (12904/58,665) of patients, while mitral valve stenosis or regurgitation was present in 76% (44520/58,665) of patients. Eleven patients (0.01%) had both aortic and mitral valve disease, and other valvular disease (ie. pulmonary, tricuspid) occurred in 2.1% (1230/58655) of patients.

There were 4 studies which included only VAF patients with mitral valve diseases (**Table 5**). Patients with moderate to severe aortic or mitral valvular disease were included in one study.^7^ These patients were specifically excluded in three of the studies. Severity of valvular disease was not specified in 8 studies. Notably, one study did exclude patients with rheumatic mitral stenosis.^14^ There were two studies that included only patients with bioprosthetic valves, while two studies listed valve replacement or repair as an exclusion criteria.^8,12,13,15^

**Table 5.** Baseline Characteristics and Complications of Patients with Mitral Valvular Disease on DOAC

| Author (Year) | patients, n | clinical follow-up, months | type of DOAC | acute ischemic stroke, n (%) | major bleeding, n (%) | intracranial bleeding, n (%) | GI Bleeding, n (%) | Fatal Bleeding, n (%) | Mortality, n (%) |
| --- | --- | --- | --- | --- | --- | --- | --- | --- | --- |
| Li (2021)<sup>10</sup> | 5833 | 12 | dabigatran, rivaroxaban | 414/5833 (7) | 359/5833 (6.2) | 74/5833 (1.3) | 285/5833 (4.9) | -- | 365/5833 (6.3) |
| Hess (2021)<sup>11</sup> | 15788 | 12 | dabigatran, rivaroxaban, apixaban, edoxaban | -- | -- | -- | -- | -- | -- |
| Guimaraes (2020)<sup>13</sup> | 500 | 12 | rivaroxaban | 3/500 (0.6) | 7/500 (1.4) | 0/500 (0) | -- | 0/500 (0) | 20/500 (4) |
| Moon (2019)<sup>14</sup> | 2792 | 12 | dabigatran, rivaroxaban, apixaban, edoxaban | 69/2792 (2.5) | 75/2792 (2.7) | 32/2792 (1.1) | 43/2792 (1.5) | -- | 161/2792 (5.8) |
| Crude Estimate (%) | 24913 | 12 (0) | -- | 486/9125 (5.3) | 441/9125 (4.8) | 106/9125 (1.1) | 328/8625 (3.8) | 0/500 (0) | 546/9125 (6) |

DOAC=direct oral anti-coagulant, GI=gastrointestinal

### Atrial Fibrillation Characteristics

Seven studies (40,439 patients) characterized the type of AF in their patients (**Table 3**). Incidental, paroxysmal, persistent, and permanent AF was found in 91% (36820/40439), 2% (795/40439), 2% (811/40439), 5% (2013/40439) of VAF patients, respectively.

### Clinical Outcomes and Complications

There were 69,741 patients observed in follow-up for a mean of 12 months (**Table 4**). Overall, AIS and systemic embolism occurred in 3.5% (1511/42877) and 0.7% (93/12599), of patients throughout the duration of the studies, respectively. Major bleeding, intracranial bleeding, and GI bleeding was present in 5.8% (3110/53953), 1.1% (534/49947), and 3.8% (1767/46802). Non-major bleeding occurred in 9.2% (335/3646) of patients. The mortality rate was 9.3% (2417/25785); fatal bleeding was present at 0.4% (16/3646).

Among patients with only mitral valve disease, at the 12 month follow-up (n=24913), 5.3% (486/9125) of patients had an AIS (**Table 5**). Major bleeding, intracranial bleeding, GI bleeding occurred in 4.8% (441/9125), 1.1% (106/9125), and 3.8% (328/8625) of patients. Fatal bleeding occurred in no patients (0/500). Overall mortality in these patients was 6% (546/9125).

Among patients on DOACs with moderate to severe mitral stenosis (single study) on dabigatran, at the 12 month follow-up, there were no reported events of major bleeding, including ICH and AIS (**Table 4**).^7^ However, three patients (5.1%) had non-major bleeding.

Among patients on DOACs who underwent bioprosthetic valve replacement or repair (2 studies), at the 12-month follow-up (n=763), AIS occurred in 8 patients (1%).^8,13^ Major bleeding, including intracranial bleeding [3/763, (0.4%)], was noted in 14 patients (1.8%), while minor bleeding occurred in 75 patients (9.8%). Mortality rate was 3.7% (28/763), and fatal bleeding occurred in one patient (0.1%) (**Table 4**).

## Discussion

In our scoping review, we identified 12 studies with 69,741 patients with VAF treated with DOAC. At the 12-month follow-up, AIS and major bleeding events occurred in 3.5% and 5.8% of patients, respectively.

DOACs for patients with NVAF have continued to demonstrate superiority or non-inferiority compared to anticoagulation with vitamin K antagonists or low-molecular-weight heparins.^19^ A meta-analysis of four pivotal trials (RE-LY, ROCKET AF, ARISTOTLE, ENGAGE AF-TIMI 48) that compared DOACS and warfarin therapy in patients with NVAF found that standard-dose DOAC users had a lower absolute risk for systemic embolism than warfarin users and lower risks of both fatal bleeding and ICH.^19^ DOACs offer the additional benefit of convenience, as there are fewer monitoring requirements, less frequent follow-up, and fewer drug and food interactions compared to vitamin K antagonist anticoagulation therapy.^1-2^

Warfarin is regarded as the first line treatment of AF in patients with valvular heart disease.^5^ Although this recommendation is primarily a result of exclusion of VAF patients in large, controlled trials that established the safety and efficacy of DOACs, post-hoc analyses of the data have shown that a significant proportion of patients in these trials did have VAF.^20^ Yet, there remains a theoretical concern that DOACs may not be as effective in preventing strokes in patients with VAF.^20^ Specifically, while NVAF is associated with a 5-fold increase in the risk of ischemic stroke, this risk is 17-fold in patients with mitral stenosis.^6^ In our review, the overall risk of AIS in patients with any type of VAF on DOAC was 3.5% (1511/42877), which is comparable to the risk in patients with NVAF on DOAC (3%) or on warfarin (3.7%) and in patients with VAF on warfarin (1.4-7%).^19^ Although, in our review, AIS seemed to occur more often in patients with mitral valvular disease (5.3%), AIS did not occur at all (0%) in a cohort of VAF patients with severe valvular disease. However, large, controlled studies are required to explain these findings, given the small number of studies that reported data sub-stratifying results based on type and severity of valve disease.

Anticoagulation therapy is associated with an increased bleeding risk, including ICH and GI bleeding.^2^ In patients with NVAF on DOAC, the risk of intracranial hemorrhage is reduced by 41-60% compared to NVAF on warfarin.^19^ In a meta-analysis, intracranial bleeding was found in 0.63% and 1.4% of NVAF patients on DOAC and warfarin, respectively.^19^ Additionally, DOAC-associated ICH have been found to be less likely to cause moderately to severely impaired consciousness (DOAC-associated ICHs: 31.3%; warfarin-associated ICHs: 39.4%) or require surgical removal (DOAC-associated ICHs: 5.3%; warfarin-associated ICHs: 9.9%).^19^ However, standard dose DOAC NVAF patients may be at a higher risk of major GI bleeding compared those on warfarin (2.54% vs 1.95%).^19^ In our review, the risk of ICH in VAF patients on DOACs was 1.1%, which is comparable to this risk in warfarin-treated VAF patients (1.1-2.5%).^19^ The risk of major GI bleeding in VAF patients on DOAC was 3.8%, similar in VAF patients on warfarin (3.6-5.3%).^22^ However, the risk of fatal bleeding in our review was found to be 0.4%, which is lower compared to in warfarin-treated VAF patients 3.9%, suggesting that a similar overall bleeding risk exists between anticoagulation therapy (ie. warfarin or DOAC) regardless of VAF or NFAF status.^23^

Overall, our results demonstrate that DOAC therapy in a heterogenous population of patients with VAF offers a comparable safety profile and efficacy compared to warfarin treated VAF with a decreased risk of fatal bleeding. However, there are a number of knowledge gaps that we identified which are crucial to address in future RCTs to establish the safety and efficacy of DOAC in VAF. Further studies are required to determine whether DOACs can adequately reduce AIS risk in VAF regardless of history of prior AIS (ie. secondary vs primary prevention), given that patients with past stroke have a higher risk of recurrent stroke (5 year recurrence rate of 12%).^24^ In our review, only 6 included studies reported on past stroke history in their study population; however, no study sub-stratified outcomes based on this baseline characteristic.

Furthermore, in our review, we found that different DOACs were used depending on study, including rivaroxaban, apixaban, edoxaban, yet rivaroxaban has been associated with higher risk of major bleeding compared with apixaban, which could affect our overall results.^25^ As such, further RCTs are required to determine which DOAC can offer the safest profile for VAF patients. Sub-stratifying data based on type of atrial fibrillation could also help guide anticoagulation decision making. Currently, there is controversy on whether the type of atrial fibrillation impacts subsequent stroke risk.^26^ Although studies have shown that patients with paroxysmal AF (PAF) have stroke risk similar to those with persistent or permanent AF, recent studies have demonstrated that PAF patients have a lower stroke risk.^26^

## Limitations

We acknowledge that there are limitations in this review. Our review was limited to primarily retrospective cohort studies. Although sample sizes in these studies were large, they did not sub-stratify or report individual data based on past medical history (ie. past stroke), type of AF, type of valvular disease, and severity of valvular disease. The studies did not indicate the amount of time patients were placed on DOACs, which can hypo or hyperinflate complication risks. The included studies did not report on all variables of interest in our review, such as HAS-BLED and incidence of AIS or major bleeding.

## Conclusion

Evidence of the safety and efficacy of DOAC anticoagulation therapy in patients with VAF is lacking. DOACs in VAF were associated with similar rates of AIS and major bleeding and decreased risk of fatal bleeding compared to warfarin therapy in these patients. Patients with mitral valve disease may be at increased risk of AIS on DOAC. However, controlled single and multi-centered studies are needed to examine complication profile and clinical outcomes data after further sub-group analysis based on AF and valvular disease type and severity.

## Data Availability

Data can be requested through the authors.

## Acknowledgements

none

